# Azithromycin alters spatial and temporal dynamics of airway microbiota in idiopathic pulmonary fibrosis

**DOI:** 10.1101/2022.09.14.22279909

**Authors:** Pieter-Jan Gijs, Cécile Daccord, Eric Bernasconi, Martin Brutsche, Christian Clarenbach, Katrin Hostettler, Sabina A. Guler, Louis Mercier, Niki Ubags, Manuela Funke-Chambour, Christophe von Garnier

**Author notes:** **Corresponding Authors:** Niki Ubags, PhD, Division of Respiratory Medicine, Department of Medicine, CHUV, Lausanne University Hospital, Lausanne, Switzerland. Shared first authorship (Pieter-Jan Gijs, Cécile Daccord). Shared last authorship (Manuela Funke-Chambour, Christophe von Garnier).

## Abstract

**Background:** High bacterial burden in lung microbiota predicts progression of idiopathic pulmonary fibrosis (IPF). Azithromycin is a macrolide antibiotic known to alter the lung microbiota in several chronic pulmonary diseases and observational studies have shown a positive effect of azithromycin on mortality and hospitalization rate in IPF. However, the effect of AZT on lung microbiota in IPF remain unknown.

**Methods:** We sought to determine the impact of a three-month course of azithromycin on lung microbiota in IPF. We assessed sputum and oropharyngeal swab specimens from 24 adults with IPF included in a randomized controlled cross-over trial of a thrice-weekly 500 mg oral azithromycin. 16S rRNA sequencing and quantitative polymerase chain reaction (qPCR) were performed to assess bacterial communities. Antibiotic resistance genes (ARG) were assessed using real-time qPCR.

**Results:** Azithromycin significantly decreased community diversity with a stronger and more persistent effect in lower airways. During treatment, turnover of airway microbiota decreased in upper and lower airways, resulting in greater similarity between microbiota of the two sites persisting one month after macrolide cessation.

Patients with increased expression of ARG had a lower bacterial load and an enrichment of the genus *Streptococcus*. In contrast, patients without increased in ARG expression had a higher bacterial load and an enrichment in *Prevotella*.

**Conclusions:** We observed that AZT caused sustained changes in the diversity and composition of the upper and lower airway microbiota in IPF, with effects on the temporal and spatial dynamics between the two sites.

## Introduction

Idiopathic pulmonary fibrosis (IPF) is a progressive and fatal interstitial lung disease of unknown origin and the most common and severe form of the idiopathic interstitial pneumonias^1,2^. Active infection plays a role in IPF progression^3^.

In healthy subjects, the composition of the microbiota in the upper respiratory tract (URT) and lower respiratory tract (LRT) has considerable similarities^4^. In the LRT, the influx of bacteria by microaspiration is counterbalanced by mucociliary clearance, resulting in a physiological turnover. Respiratory health is therefore associated with a dynamic turnover of the microbiota between the URT and the LRT.

Converging evidence suggests that in IPF the spatial and temporal dynamics of the airway microbiota are disturbed. LRT microbiota is more abundant^5,6^ and less diverse^7^, suggesting either greater local growth, accumulation due to impaired clearance, or both (**Figure 1**). One consequence is greater dissimilarity between URT and LRT microbiota, with LRT microbiota carrying a majority of genes absent in the URT^8^. This, alongside with evidence from longitudinal data on persistence of LRT microbiota disturbances^9^, provides additional evidence for a lower microbiota turnover in IPF.

**Figure 1.**
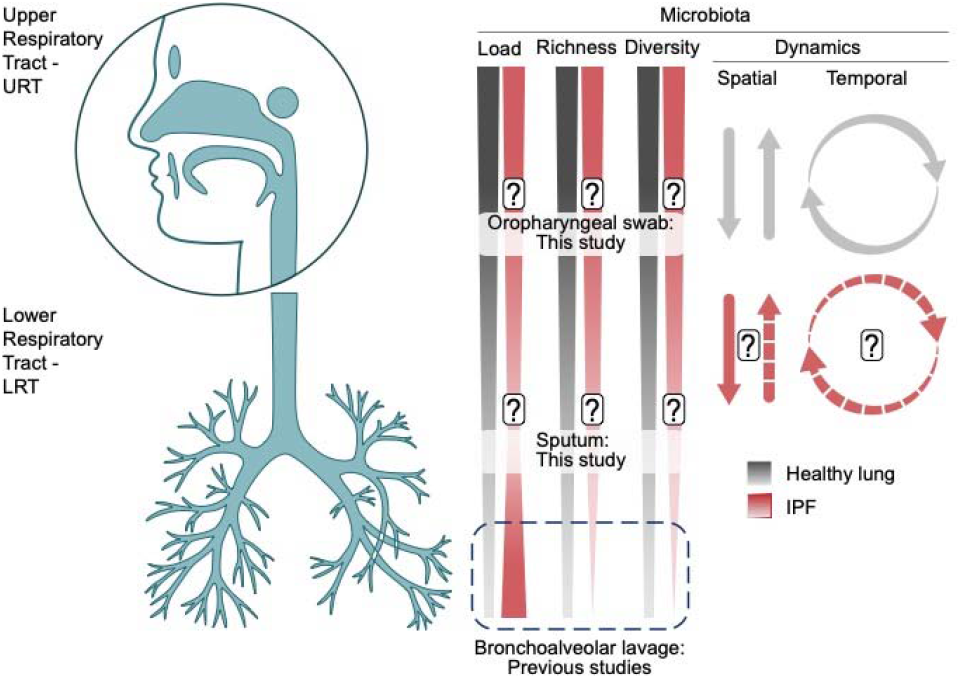
In health, the LRT microbiota is a community of transiently present microorganisms originating from the URT, whereas in IPF the LRT microbiota is a thriving community with a higher bacterial load and a decreased community diversity. The spatial dynamics between the URT and LRT microbiota in IPF, their turnover rates, and how these are affected by AZT remains unknown. *Abbreviations:* LRT = lower respiratory tract; URT = upper respiratory tract; IPF = idiopathic pulmonary fibrosis; AZT = azithromycin

Preclinical data suggest that disruption of LRT microbiota precedes chronic lung epithelial injury and repair^7,10^, potentially perpetuating inflammation^9^. Bacterial load is currently the LRT microbiota feature most consistently associated with disease, in terms of progression, exacerbations and mortality. Bacterial load is higher in IPF patients than in healthy subjects, making it a potential therapeutic target^11^.

A single-centre retrospective study showed good tolerance to prophylactic AZT in IPF patients, with fewer non-elective hospitalizations^12^. Also, treatment with AZT during acute IPF exacerbations improved survival rates compared to fluoroquinolones in a retrospective single-center study^13^. The impact of AZT on the respiratory microbiota in IPF is unknown. In patients with severe asthma and emphysema, multiple effects on the composition and structure of the LRT microbiota, as well as increased anti-inflammatory bacterial metabolites have been reported^14,15^. This effect of AZT on LRT microbiota was independent of a decrease in bacterial load. Acquired macrolide resistance is a global health concern and macrolide administration increases the carriage of macrolide-resistant bacteria in URT^16,17^ and LRT^18^. Higher carriage was observed after AZT for seven genes involved in antibiotic resistance, five of which associated with macrolide resistance, and two with tetracycline resistance^14^. Additionally, the airway resistome positively correlates with bacterial load in chronic obstructive pulmonary disease (COPD)^19^.

This study examines the impact of a three-month treatment of AZT on the airway microbiota of IPF patients.

## Materials and Methods

### Study population and sample collection

This study is a post hoc analysis of samples collected in the “*Azithromycin for the Treatment of Chronic Cough in Idiopathic Pulmonary Fibrosis”* study (Clinical trial identifier: NCT02173145)^20^. This previous study was a multi-centre, double-blind, randomised, placebo-controlled cross-over trial to determine the effect of AZT on chronic cough in IPF patients (see Supplementary materials for key inclusion and exclusion criteria).

Patients underwent a 12-week treatment period with oral AZT 500 mg 3 times per week, and a 12-week treatment period with placebo 3 times per week in random order. The two periods were separated by a 4-week washout period.

Expectorated sputum and oropharyngeal swab (OPS) samples were collected both before and after AZT and placebo periods, and an additional specimen was collected after the 4-week washout period following the second treatment period. Samples were stored at -80°C until analysis. The study design and detailed sample collection are presented in Figure E1 in the Supplementary data.

### DNA extraction and 16S rRNA amplicon quantification

Sputum and OPS samples were treated with dithiothreitol to homogenise the mucosal phase and DNA was extracted using the DNeasy UltraClean microbial kit (Qiagen, Hilden, Germany), with inclusion of a lysozyme digestion step (see details in Supplementary data).

Negative controls (N = 11) underwent the same procedure and included blank swabs, blank sputum collection tubes, as well as blank DNA extractions (reagent control).

To obtain a proxy of bacterial load, we determined the copy numbers of the 16S rRNA gene by qPCR using previously reported primers specific to panbacteria^21^ (see **Supplementary Table E1** for full sequences). Standard curves were obtained using purified amplicon products.

### Bacterial 16S rRNA amplicon sequencing

Amplicon sequencing targeted the V1-V2 region of the 16S rRNA gene with primers F-27 and R-338 (see Supplementary Table E1 for full sequences and Supplementary Data for details). Amplification was performed using the Accuprime Taq DNA Polymerase High Fidelity kit (Invitrogen, Waltham, MA). No-template PCR reaction controls (N = 2) were included. Libraries were loaded onto an Illumina MiSeq using pairwise chemistry, generating 250 × 2 read lengths (Lausanne Genomic technologies facility, University of Lausanne, Switzerland).

### Analysis of antibiotic resistance gene carriage

Quantification of antibiotic resistance genes (ARG) carriage targeting 23S ribosomal RNA methyltransferases (*erm*(B) and *erm*(F)), ATP-binding cassette ribosomal protection protein (*mel* and *msr*[E]), major facilitator superfamily antibiotic efflux pump (*mef*), and tetracycline-resistant ribosomal protection proteins (*tet*[M] and *tet*[W]) was performed on sputum specimens using dye-based (SsoAdvanced Universal SYBR Green, Bio-Rad) or probe-based real-time PCR assays, using primer pairs, probes and conditions previously described^14^. The copy numbers of resistance genes per sample were normalised relative to the copy numbers of the 16S rRNA gene. To obtain a synthetic picture of ARG carriage per sample, the counts obtained for each individual gene were scaled from 0 to 1 to provide equal importance to each gene, and the cumulative counts were reported.

### Bioinformatics and statistical analysis

All analyses were performed in R version 4.1.0^22^. Demultiplexing, removal of chimeric and short reads, single-base resolution of reads into amplicon sequence variants (ASVs) using the Divisive Amplicon Denoising Algorithm 2 (DADA2) algorithm^23^, and taxonomic annotation using the SILVA database^24^ were performed using a dedicated pipeline available at https://github.com/chuvpne/dada2-pipeline. Analyses were performed on a rarefied dataset at a sequencing depth of 11,741 (**Figure E2;** see Supplementary materials for details on processing and quality control).

Multiple group comparisons were made using Kruskal Wallis test with Dunn’s post hoc test and Holm’s adjustment. The Wilcoxon signed-rank test was used to compare paired data. In all tests, we considered an alpha significance level of 0.05.

To assess the difference in bacterial community composition between OPS and sputum samples during each phase of AZT treatment, we computed the Aitchison distance, corresponding to the Euclidean distance between the samples after centered log-ratio transformation^25^.

Comparisons of proportions of ASVs represented only in one condition or shared between two conditions were assessed using the chi-square goodness-of-fit test.

To test for differences in microbiota composition between the different phases of AZT treatment, we used a matrix built on unweighted UniFrac distance and conducted a Permutational Multivariate Analysis of Variance (PERMANOVA) with 999 permutations.

## Results

### Study cohort

Of the 25 patients who completed randomization, 12 were initially randomized to intervention and 13 to placebo. 20 patients completed the study. The Consolidated Standards of Reporting Trials diagram is shown in **Figure E3** in Supplementary materials.

24 patients had at least one specimen, with a total of 67 sputum and 90 OPS specimens. **Table 1** shows patient demographics and clinical characteristics.

**Table 1.** Demographics and clinical characteristics of study cohort^20^ *Abbreviations*: UIP = usual interstitial pneumonia; HRCT = high-resolution computed tomography; TLC = total lung capacity; FVC = forced vital capacity; FEV1 = forced vital capacity in 1 second; DLCO = diffusing capacity of the lung for carbon monoxide. Values are presented as frequency and percentage or mean and standard deviation (SD)

| Characteristics | Study cohort (N=25) |
| --- | --- |
| Age, years | 67 (8) |
| Sex, men | 23 (92%) |
| Previous smokers | 17 (68%) |
| Current smoker | 1 (4%) |
| **Diagnosis** |  |
| Definite UIP pattern on HRCT | 18 (72%) |
| Surgical lung biopsy available | 12 (48%) |
| Multidisciplinary team discussion performed | 19 (76%) |
| **Treatment** |  |
| Pirfenidone | 9 (36%) |
| Nintedanib | 11 (44%) |
| Proton pump inhibitor | 13 (52%) |
| Oxygen therapy | 9 (36%) |
| **Pulmonary function tests** |  |
| TLC, % predicted | 60 (12) |
| FVC, % predicted | 65 (16) |
| FEV1/FVC, % | 86 (7) |
| DL<sub>CO</sub>, % predicted | 43 (16) |
| **Comorbidities** |  |
| Chronic rhinitis | 8 (32%) |
| Sinusitis | 2 (8%) |
| Gastroesophageal reflux disease | 4 (16%) |
| Cardiac disease | 11 (44%) |
| Pulmonary hypertension | 3 (12%) |
| Diabetes | 4 (16%) |

Bacterial quantification, 16S rRNA gene sequencing and ARG analysis were performed on all specimens, except on 1 OPS specimen with insufficient DNA. 10 OPS specimens not meeting the 11’741 read threshold were excluded from further sequencing analysis (**Figure E4** in Supplementary materials). Finally, 67 sputum and 79 OPS specimens were analyzed.

To investigate temporal changes in respiratory microbiota linked to AZT, we distinguished five treatment phases: *PreAZT* for specimens collected before the start of AZT treatment, available only in patients who started with placebo (N=35; sputum=14; OPS=21), *StartAZT* for specimens collected at the beginning of AZT (N=31; sputum=15; OPS=16), *EndAZT* for specimens collected at the end of AZT (N=23; sputum=12; OPS=11), *PostAZT_1 month* for specimens collected 1 month after the end of treatment (N=32; sputum=14; OPS=18) and *PostAZT>=3 months* for specimens collected 3 months or more after the end of AZT (N=25; sputum=12; OPS=13).

### The respiratory microbiota of IPF patients is distinct from environmental noise

As our study used low volume specimens from body sites with low microbial biomass, it was vulnerable to environmental noise^26^. Nevertheless, we detected a markedly higher load in both LRT (*p* < 0.001; **Figure E5a**) and URT (*p* < 0.001; **Figure E5b**) specimens compared to procedural controls. Rank abundance analysis further showed that the dominant bacteria were mostly different in patient samples versus controls (**Figure E5c**) and allowed us to exclude from further analysis two ASVs dominant in controls (ASV6_*Cutibacterium*; ASV26_*Pseudomonas*).

### The upper and lower airway microbiota of IPF patients is altered after AZT treatment

There was no difference in bacterial load between the different treatment phases in either LRT (*p* = 0.95) or URT (*p* = 0.22; **Figure 2**), but we observed a decrease in community richness after AZT, with a stronger and more persistent effect in LRT (*p* = <0.001) compared to URT (*p* = 0.008; **Figure 3a**). A decrease in bacterial phylogenetic diversity was also observed in both LRT (*p* = <0.001) and URT (*p* = 0.0093), without reverting to the pre-treatment level until the end of the observation period for LRT and more transiently for URT (**Figure 3b**). In contrast, evenness increased transiently after treatment in LRT only (*p* = 0.0021; **Figure 3c**). The Shannon diversity index reflects the effects of the AZT on richness (Figure 3d). In addition, we observed a transient increase in the dominance of core community ASVs in LRT (*p* = 0.014), with a non-significant trend in URT (*p* = 0.22; **Figure E6** Principal coordinate analysis (PCoA) based on unweighted UniFrac distance, which accounts for phylogeny showed a significant shift between pre- and post-treatment communities, both in LRT (*p* = 0.008; **Figure E7a**) and URT (*p* = 0.009; **Figure E7b**).

**Figure 2.**
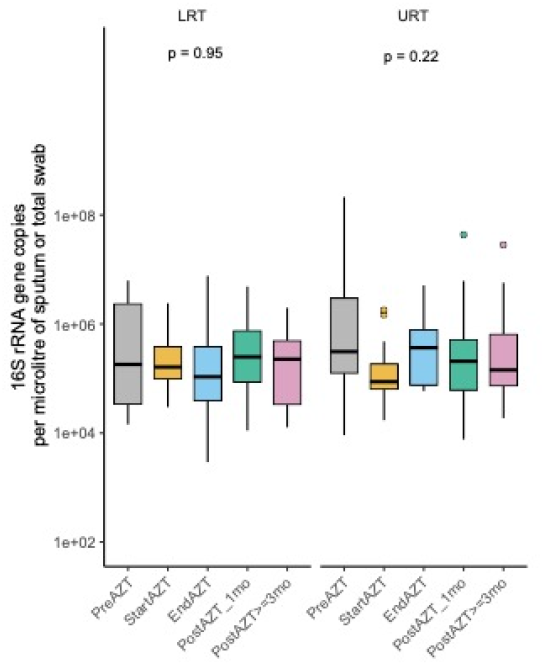
qPCR determination of 16S rRNA gene copy number per microlitre of sputum or total OPS samples showing no variation in bacterial load during the different phases of the study. *Abbreviations:* qPCR = quantitative polymerase chain reaction; OPS = oropharyngeal swab

**Figure 3.**
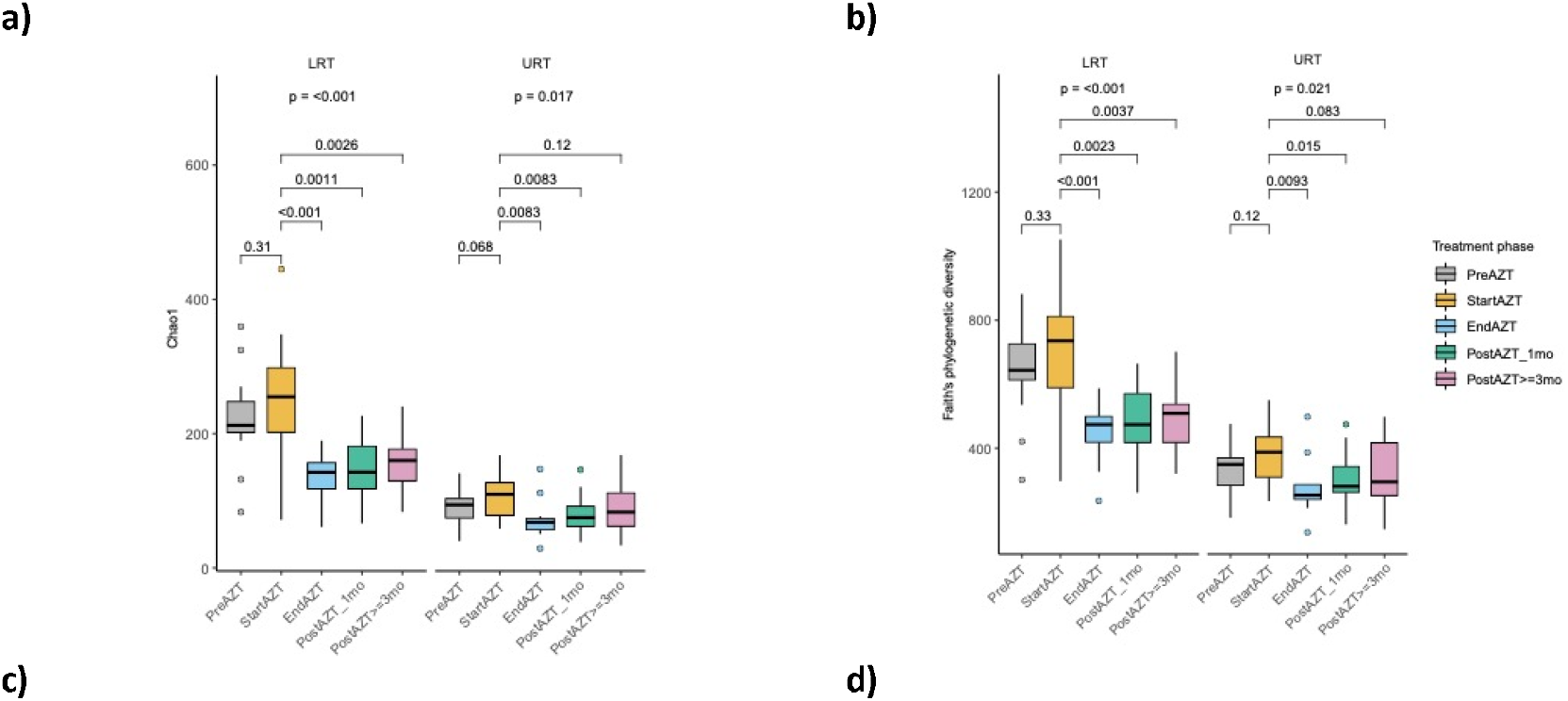
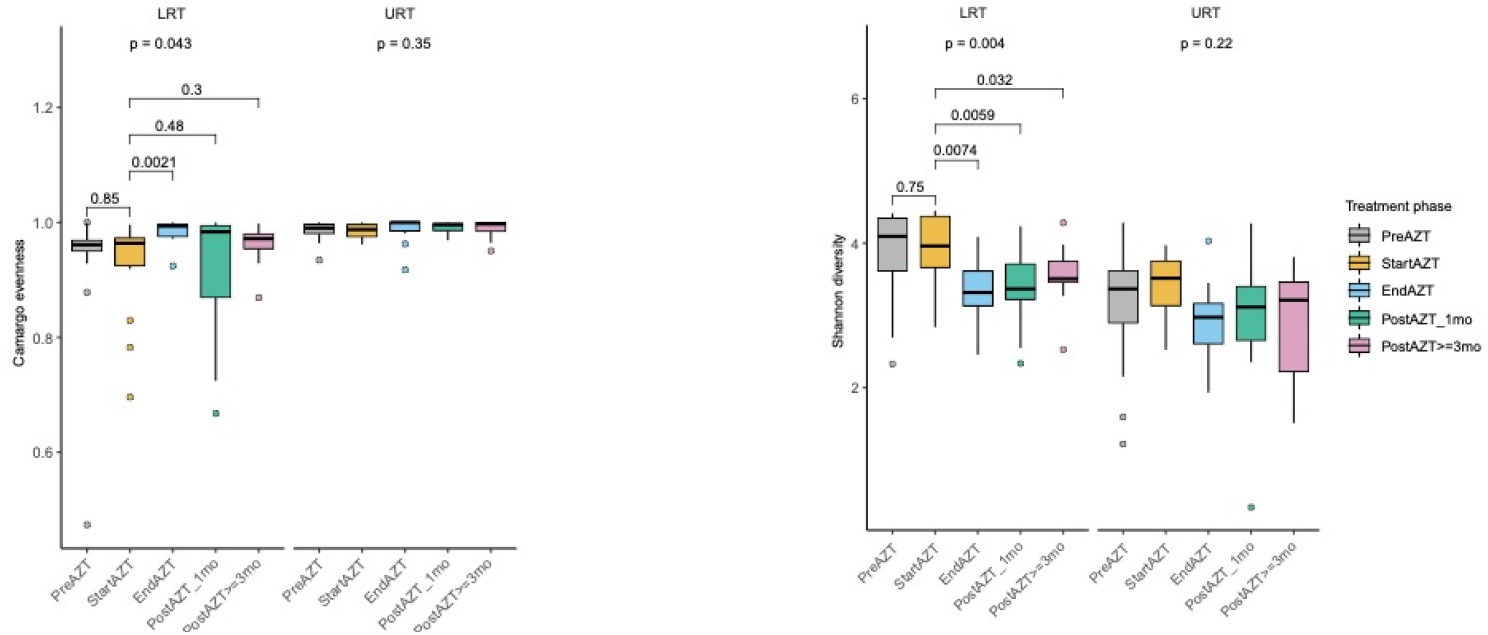
Decrease in community diversity after AZT treatment. (*a* to *d*) Alpha diversity metrics showing a decrease in Chao1 richness (*a*) and Faith’s diversity (*b*) during AZT treatment in LRT and URT, an increase in Camargo evenness in LRT (*c*), and a decrease in Shannon diversity in LRT (*d*). *Abbreviations*: AZT = azithromycin; LRT = lower respiratory tract; URT = upper respiratory tract

Together, these results show a broad impact of AZT on the respiratory microbiota of IPF patients.

### AZT treatment reduces spatial dissimilarity between URT and LRT microbiota and affects their temporal dynamics

Based on unweighted UniFrac distance, bacterial communities in URT and LRT were distinct (**Figure 4a**). Per-patient comparisons showed greater similarity between URT and LRT samples at the end of AZT treatment, with the effect persisting one month after treatment with subsequent attenuation thereafter (**Figure 4b**). Divergence of the microbiota between the two sites was driven by ASVs present only in LRT, more numerous (*p* < 0.001) than those present only in URT or shared between the two sites, in each treatment phase. The transient decrease in the divergence between URT and LRT microbiota after AZT was associated with a non-significant decrease in the proportion of ASVs represented in LRT only, and a marginal increase in the number of ASVs shared by both sites (**Figure 4c**).

**Figure 4.**
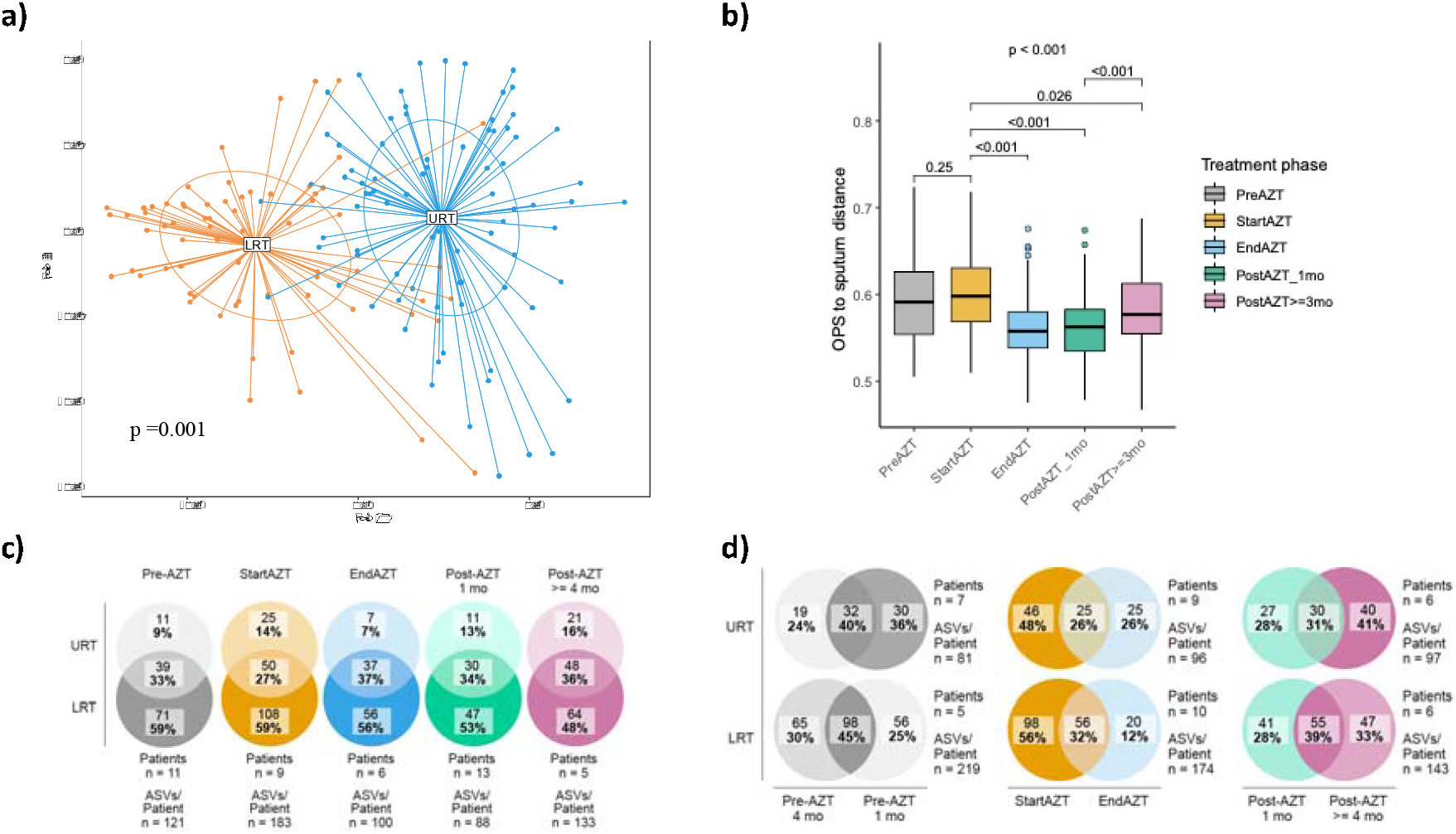
AZT treatment reduces dissimilarly between LRT and URT microbiota. (*a*) PCoA based on unweighted UniFrac distance showing an overall difference in bacterial community composition between URT and LRT. (*b*) Aitchison distance showing a decrease in dissimilarity between URT and LRT microbiota between the start and end of treatment. For each treatment phase, Aitchison distance was calculated between each OPS sample and the centroid obtained from the corresponding set of sputum samples. (*c*) Venn diagrams of the numbers and proportions of ASVs represented in either the LRT or URT alone, or in both sites, showing that the decrease in dissimilarity between the LRT and URT microbiota during treatment was in part driven by a decrease in the proportion of ASVs present in the LRT alone.(*d*) Venn diagrams of the numbers and proportions of ASVs represented either at the start, at the end or retained over a 3-month time window, showing a decrease in ASV turnover after AZT treatment in both LRT and URT. *Abbreviations*: AZT = azithromycin; LRT = lower respiratory tract; URT = upper respiratory tract; PCoA = principal coordinate analysis; OPS = oropharyngeal swab; ASVs = amplicon sequence variants

Analysis of samples from IPF patients first receiving placebo allowed quantification of bacterial turnover over a three-month period in the absence of AZT. We found a mean of 40% ASVs retained in URT, and 45% in LRT (**Figure 4d, left diagrams**).

During AZT treatment, patients had a slower turnover in airway microbiota, resulting in fewer ASVs present at the end compared to the start of AZT treatment in LRT (*p* < 0.001) and URT (*p* = 0.008). In LRT, the number of newly acquired ASVs was also lower than the number of retained ASVs (*p* = 0.004), in contrast to the observations in patients receiving placebo first (**Figure 4d, middle diagrams**).

However, the bacterial turnover after treatment was increased compared to the placebo first group, with numbers of newly acquired ASVs exceeding those of cleared ASVs in both the LRT (*p* = 0.045) and URT (*p* = 0.026; **Figure 4d, right diagrams**).

We observed that bacteria cleared from LRT during treatment, which accounted for 13 to 47% of the local community at the start of treatment, were already present in this site (between 7 and 24% relative abundance) at least four months before treatment started. The effect of treatment on these bacteria was long-lasting, with only partial resilience five months after the end of treatment. These same bacteria followed similar kinetics in URT, with the relative abundance ranging from 2-42% in the 4 months prior to the start of treatment and then decreasing to less than 0.05% (3 patients) or below detection (3 patients) (**Figure 5a**).

**Figure 5.**
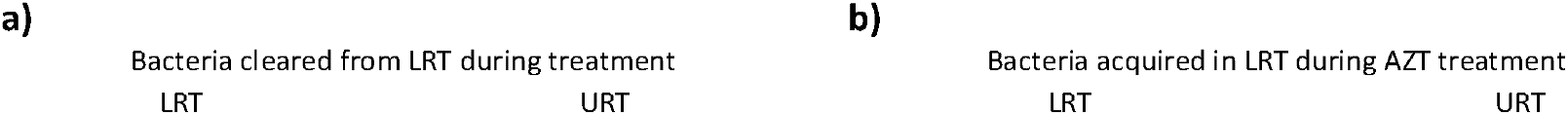
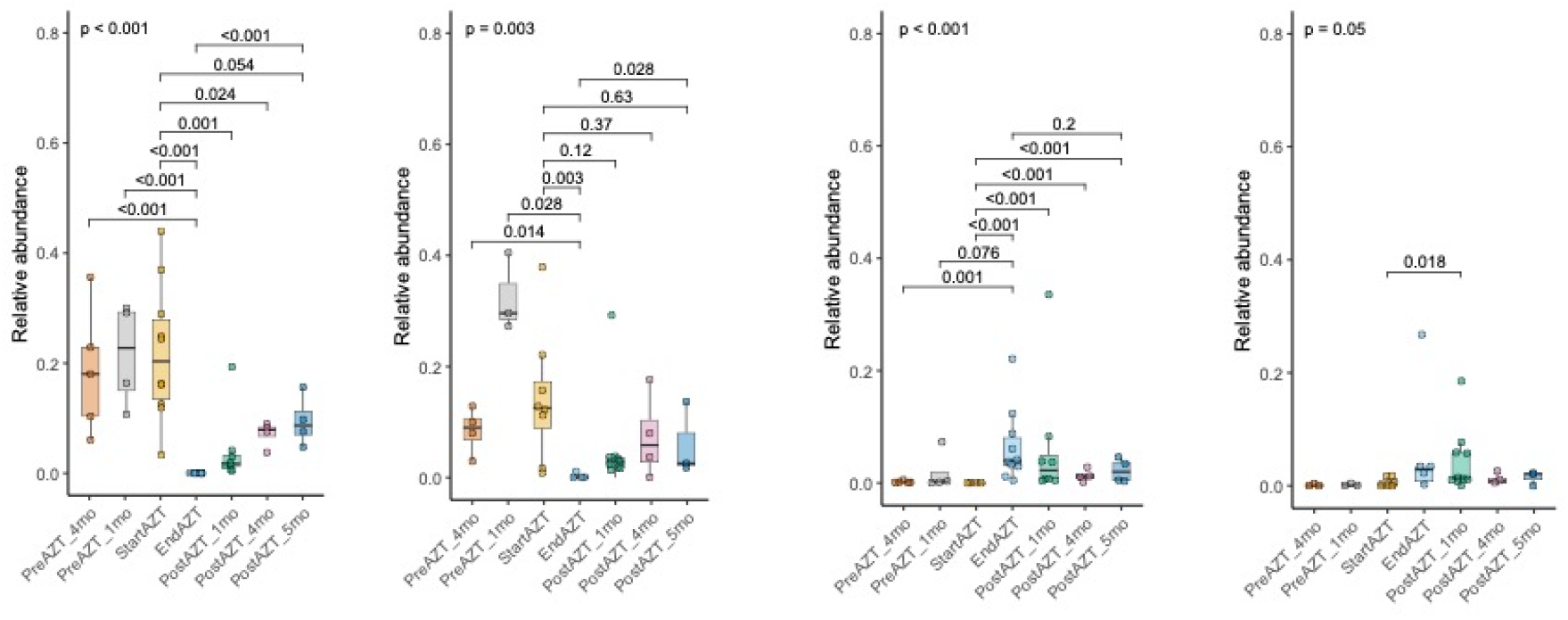
Longitudinal changes in the relative abundance of bacteria cleared (*a*) or acquired (*b*) in the LRT during AZT treatment, showing different magnitudes of change in relative abundance between cleared and acquired bacteria, and similar kinetics between LRT and URT. *Abbreviations:* LRT = lower respiratory tract; AZT = azithromycin; URT = upper respiratory tract

Bacteria acquired in LRT during treatment were represented in some cases at low levels during the four months prior to treatment. Their local relative abundance was highest at the end of treatment, without reaching the levels of bacteria cleared by the treatment, before decreasing during the 5 months following the end of treatment. These same bacteria were also poorly represented in URT prior to treatment (less than 1% in 7 of the 8 patients with data available). Their local relative abundance also increased during treatment, and, for some patients, these bacteria remained detectable in URT for up to 5 months after the end of treatment (**Figure 5b**).

Taken together, these observations indicate marked changes in the spatial distribution and temporal dynamics of the airway microbiota of IPF patients secondary to AZT treatment.

### Changes in the composition of LRT microbiota correlate with antibiotic resistance gene carriage

We next investigated whether alterations in respiratory microbiota during AZT treatment were related to ARG acquisition, focusing on LRT and previously described target genes^14^. We observed that ARG carriage was limited to a minority of LRT samples and ASVs (**Figure 6a)** and that almost all samples with ARG were collected at the end of AZT treatment or later (**Figure 6b**). This was confirmed by longitudinal within-patient analysis, which showed a peak in total ARG carriage at the end of AZT treatment, with substantial inter-individual differences in post-treatment kinetics (**Figure 6c**).

**Figure 6.**
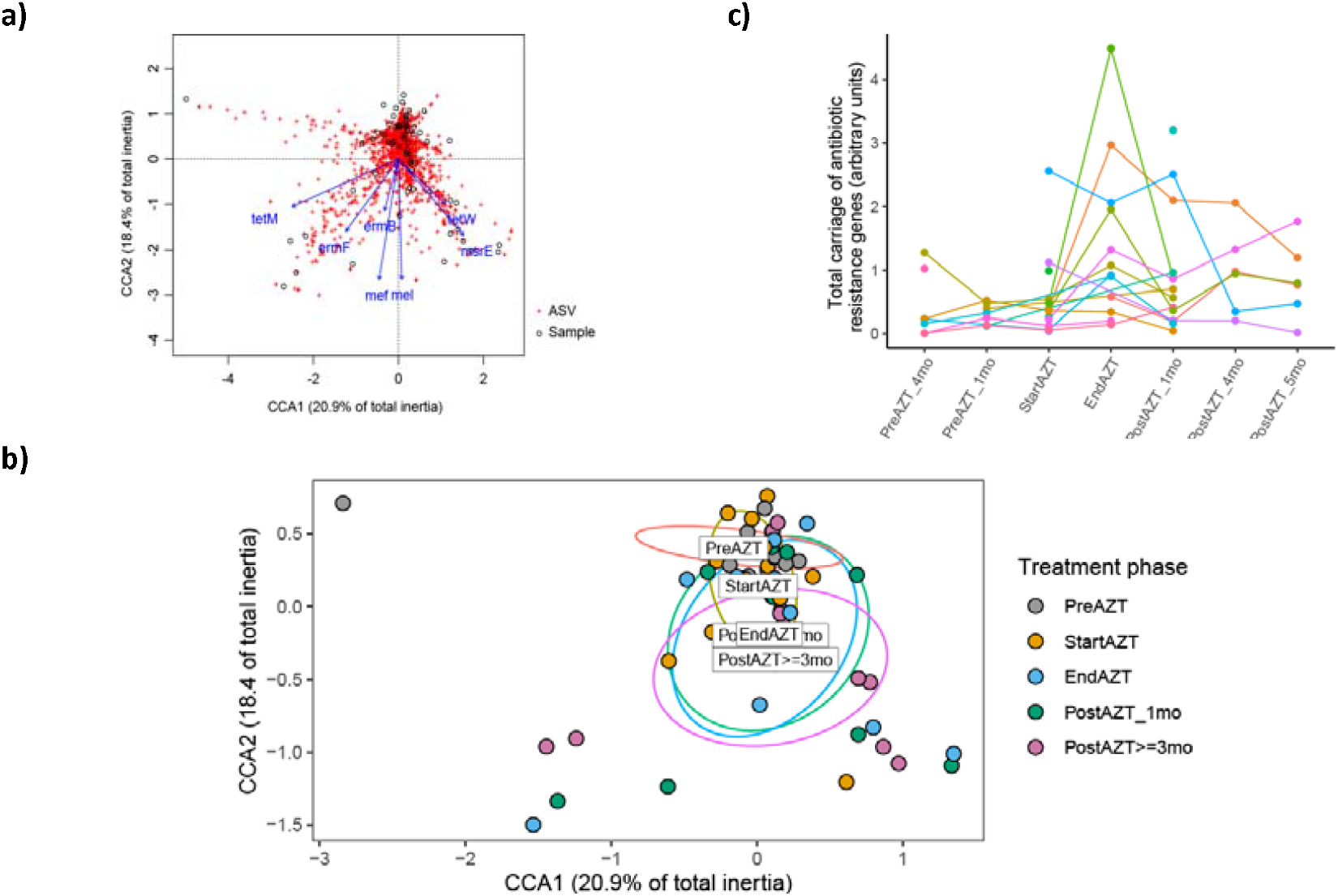
Acquisition of ARG during AZT treatment in LRT. (*a* and *b*) Canonical correlation analysis showing that ARG carriage was limited to a minority of sputum samples (circles) and ASVs (crosses) (*a*), and stratification by treatment phase showing that almost all samples with ARG were taken at the end of AZT treatment or later (*b*). (*c*) Within-patient monitoring of the cumulative sum of normalised copy numbers of the full set of ARG genes, focusing on patients with more than one sputum sample available, showing the increase in ARG carriage after the start of AZT treatment. *Abbreviations:* ARG = antibiotic resistance genes; AZT = azithromycin; LRT = lower respiratory tract; ASVs = amplicon sequence variants

To investigate the implications for LRT microbiota of an increase in ARG carriage, we separated patients with LRT samples available at the start and end of AZT treatment (N = 10) into two groups of five patients according to the median change in the cumulative sum of seven pooled ARG (median fold-change = 3.8) (**Figure E8**).

This revealed an association between ARG carriage and bacterial load, with a higher load in patients with stable resistance compared with those with increased resistance during AZT treatment (*p* < 0.001) (**Figure 7a**). However, there was no change in bacterial load between the different treatment phases, whether ARG carriage status was stable or increased (**Figure E9**).

**Figure 7.**
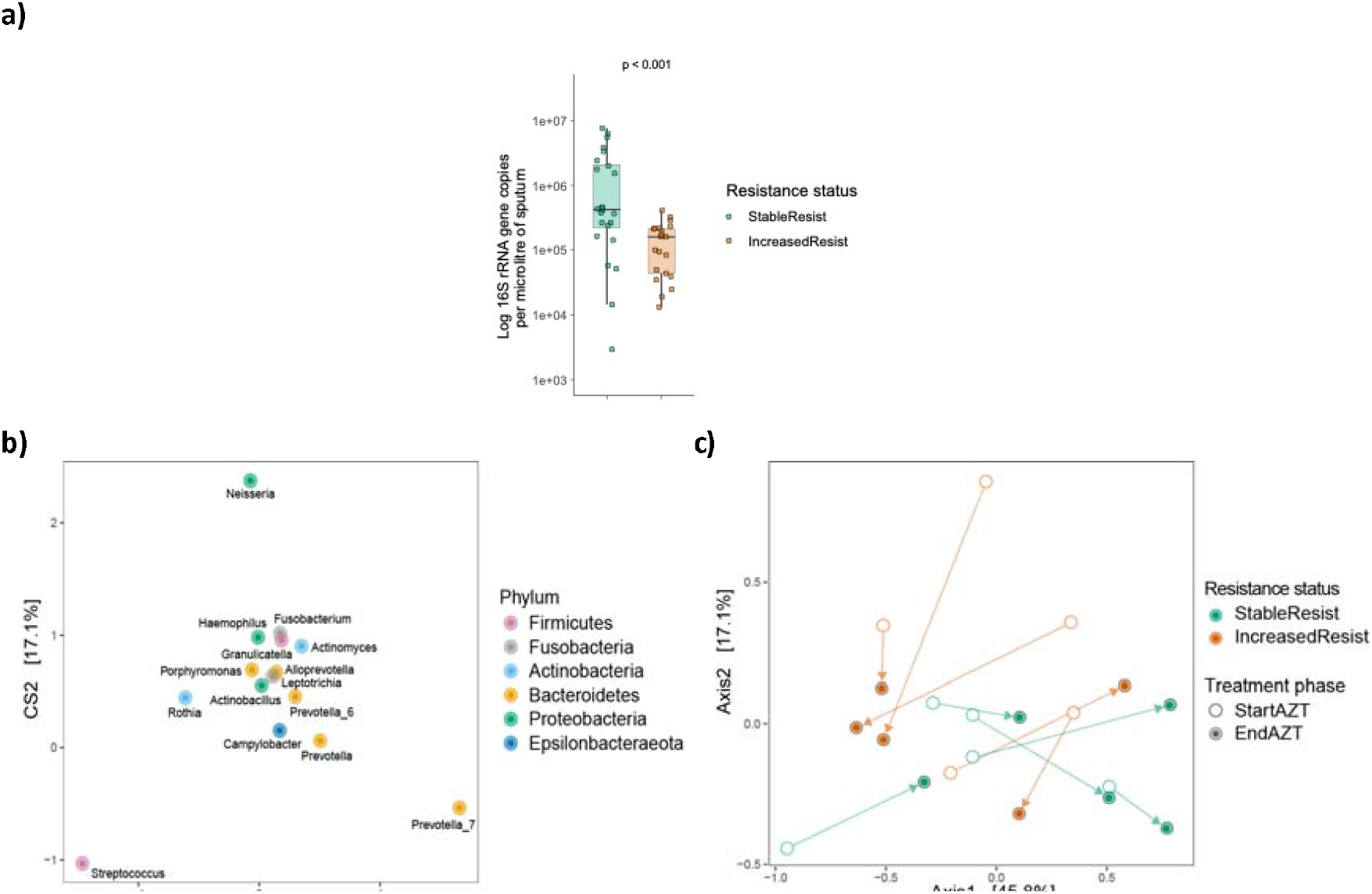
Relationship between the difference in ARG carriage during AZT treatment, bacterial load and the 15 dominant genera of the LRT microbiota. (a) qPCR determination of bacterial load showing lower levels in patients with increased resistance. (b and c) Double PCoA showing the influence of *Prevotella_7* abundance in the compositional changes occurring during AZT treatment in patients with stable resistance, respectively *Streptococcus* abundance in patients with increased resistance. In panel c, the arrows link LRT samples collected from a single patient at the start and end of AZT treatment. *Abbreviations:* ARG = antibiotic resistance genes; AZT = azithromycin; LRT = lower respiratory tract; qPCR = quantitative polymerase chain reaction; PCoA = principal coordinate analysis

We observed changes in the composition of LRT microbiota between the start and end of treatment, depending on resistance status. Specifically, double PCoA focused on the 15 most abundant genera showed that AZT treatment was associated with a shift in composition influenced by *Prevotella_7* abundance, in all patients with stable resistance and only in one patient with increased resistance. In contrast, the other four patients with increased ARG carriage showed compositional changes mainly driven by *Streptococcus* (**Figure 7b and c**).

Genus abundance analysis during treatment confirmed an increase in *Prevotella*_7 and a decrease in *Streptococcus* in patients with stable resistance, and an increase in *Streptococcus* in 4 out of the 5 patients with increased resistance (**Figure E10a and b**). Finally, enrichment of *Prevotella*_7 in patients with stable resistance during treatment (*p* = 0.006; **Figure E10c**), and of *Streptococcus* in those with increased resistance (*p* = 0.002; **Figure E10d**), was also observed when all samples of these patients were considered, including those taken before the start or after the end of treatment.

Together, these observations link the resistome to the composition of LRT microbiota, with a direct correlation with local bacterial load, but without change in lung function during the study period (**Figure E11**)^28^

## Discussion

This study provides valuable insight on the spatial and temporal distribution of the respiratory microbiota in IPF by comparing URT and LRT, microbial turnover, and the impact of AZT on these dynamics.

Our findings suggest an attenuating effect of AZT on airway ecological disruption but also disturbance in LRT through a decrease in community diversity and an increase of potential pathogenic community members. These effects persisted up to five months after the end of treatment, possibly due to AZT retention in lung macrophages^27^. No changes in clinical status were observed during the study period^20^.

URT and LRT communities in our cohort of IPF patients differed significantly, as in other chronic respiratory diseases^28,29^, with microbial richness in LRT exceeding that in URT, which contrasts with the situation in healthy subjects.

Greater decrease in microbial richness in LRT versus URT during AZT treatment, paralleled by a diminishing phylogenetic dissimilarity between microbiota of the two sites, suggests that treatment decreases airway ecological disturbance. The absence of a concomitant decrease in bacterial load implies that the numerous taxa cleared were replaced by a smaller number of taxa already present and becoming more dominant. As increased dominance may alter the bacterial impact on host, it is often interpreted as being harmful, but little is known about the threshold for detrimental dominance, which depends on bacteria and the clinical context.

Further evidence of AZT treatment effects was investigated by assessing bacterial temporal dynamics. In the absence of AZT, bacterial turnover was lower in the LRT compared to the URT. Combined with excessive microbial richness in LRT, this suggests a local persistence of bacteria potentially harmful to the host. AZT treatment had two effects on this turnover, namely by eliminating taxa (decrease in richness) and by preventing the acquisition of new taxa. However, these effects vanished progressively during the months following treatment.

The shift in LRT microbiota composition associated with AZT, and the decrease in richness and phylogenetic diversity, mirrors the findings reported in asthma and emphysema^14,15^. These reported alterations were mainly driven by a mutually exclusive increase in relative abundance of the two predominant genera of LRT microbiota in IPF, *Streptococcus* (Gram-positive *Firmicutes*) and *Prevotella* (Gram-negative *Bacteroidetes*). Enrichment of these genera was consistent with the development of macrolide resistance previously described in cystic fibrosis^30,31^. The difference we observed with the increase in ARG carriage during AZT treatment when either of these genera predominated suggests that different mechanisms may be present. The increase in ARG carriage (between 0.5- and 11-fold for 12 weeks of AZT exposure) across the cohort was consistent with findings reported in asthma^14^. While *Streptococcus* enrichment was associated with a marked increase in ARG carriage (in 4 out of 4 patients), *Prevotella* enrichment was linked to more stable ARG carriage (in 5 out of 6 patients). *Prevotella* acquisition of AZT resistance could be due to carriage of genes not listed in our targets, although we included the most likely candidates based on a previous metagenomic study of AZT treatment in severe asthma^14^.

A larger sample size will be necessary to strengthen conclusions and to perform in-depth analysis to determine whether carriage of genes involved in tetracycline versus macrolide resistance is associated with different bacteria.

Although patients remained clinically stable during the study period, it is difficult to predict the impact of even a relatively transient predominance of *Streptococcus* or *Prevotella* on lung health. Both genera include commensal species that can act as opportunistic pathogens under permissive conditions. Some members of *Streptococcus* have been associated with IPF progression^6^. *Prevotella* are maintained at low levels in the healthy lung, where they establish a subclinical level of inflammation favoring immune surveillance^4^. However, their abundance is increased in IPF where they can become predominant^32^. Furthermore, *Prevotella* are involved in several inflammatory diseases^33^, including periodontitis, with a possible impact on the frequency of exacerbations in COPD^34^. Accordingly, preclinical evidence suggests that *Prevotella* predominance in the airways promotes pulmonary fibrosis through a mechanism involving IL-17B^35^. Decreased microbial diversity in IPF, such as observed with the predominance of *Streptococcus* or *Prevotella*, has previously been associated with increased alveolar concentrations of pro-inflammatory cytokines and profibrotic growth factors^10^.

Further studies are required to determine the relationship between alterations of microbiota and gene expression in the airways during AZT treatment.

Our study has limitations. First, we report an association between AZT and ARG carriage, but separate analysis of microbiota by amplicon sequencing and ARG carriage by qPCR does not allow us to identify bacteria that acquire resistance during treatment. Genome-wide metagenomic analyses would answer this question. Second, the absence of a healthy control group prevented us from assessing whether disruption of the airway microbiota by AZT is specific for IPF. Third, although active infection or antibiotic therapy 4 weeks prior to enrolment was an exclusion criterion for the study, a preconditioning of the respiratory microbiota that might have impacted the subsequent AZT effect in some patients cannot be excluded.

In conclusion, we found that AZT alters the spatial and temporal dynamics of airway microbiota in IPF patients with a decline in richness and phylogenetic diversity in URT and LRT, without impact on bacterial load. The AZT impact was characterized by increased *Prevotella* or *Streptococcus* abundance and persisted with partial resilience five months after treatment. Also, increased ARG carriage was present in half of patients in the presence of *Streptococcus* predominance. This study provides novel insights into airway ecology disturbances in IPF by longitudinal sampling of URT and LRT and expands on previous knowledge from studies in distal LRT. From a clinical care perspective this approach also encourages non-invasive sampling of more than one respiratory tract site, particularly in patients with IPF for whom bronchoscopic sampling is frequently associated with an unfavorable risk-benefit ratio.

## Data Availability

All data produced in the present study are available upon reasonable request to the authors

https://github.com/chuvpne/dada2-pipeline.

https://github.com/CHUVpulmonology/Airway_microbiota-Lung_fibrosis-Azithromycin

## Acknowledgements

We thank the patients who participated in the study and the study nurses from Bern, Basel, St. Gallen and Zurich for their work and data acquisition. We also thank the Lung Association of the Canton of Vaud (Ligue Pulmonaire Vaudoise, LPV) and the Research Fund of the Swiss Lung Association, Bern for financial support. 16S rRNA sequencing was performed at the Lausanne Genomic Technologies Facility, University of Lausanne, Switzerland (https://www.unil.ch/gtf/en/home.html).

## Notes

### Competing Interest Statement

All authors have completed the ICMJE uniform disclosure form at www.icmje.org/coi_disclosure.pdf and declare: no support from any organisation for the submitted work; CC has received honorariums from GSK, Novartis, Vifor, Boehringer, Astra Zeneca, Sanofi and does consulting for Daiichi Sankyo, MFC has received honorariums for MSD, Novartis and received consulting fees from Boehringer Ingelheim and Daiichi Sankyo

### Clinical Trial

NCT02173145

### Funding Statement

This study was supported by a fund from the Lung Association of the Canton de Vaud (Ligue Pulmonaire Vaudoise, LPV), Switzerland, awarded to CD and EB. The initial interventional which allowed sample availability was supported by the Research Fund of the Swiss Lung Association (Lungenliga Schweiz), awarded to MFC.

### Author Declarations

Ethical approval was obtained by the Swiss Ethics Committee,Bern (KEK 002/14)before the start of the study, and all patients provided written informed consent before inclusion

