## Supplemental Material for "Azithromycin alters spatial and temporal dynamics of airway microbiota in idiopathic pulmonary fibrosis"

* Shared first authorship

^+^ Shared last authorship

**Corresponding Author**:

Niki Ubags

Division of Respiratory Medicine, Department of Medicine, CHUV, Lausanne University Hospital, Lausanne, Switzerland

**Supplementary materials and methods**

Study population

The study was conducted between August 2014 and August 2019; Key inclusion criteria were age over 18 years and a diagnosis of IPF according to current diagnostic guidelines^2^. Ethical approval was obtained prior to the start of the study (KEK 002/14), and all patients provided written consent prior to inclusion in the study. Patients with one or more of the following criteria were excluded : any change in medication or respiratory infection within four weeks prior to inclusion, known allergy or intolerance to macrolide antibiotics, known cardiac arrhythmia, severe renal failure, history of hepatitis, current alcohol or drug abuse, serum bilirubin level of > 50 µmol/L, elevated aspartate transaminase or alanine transaminase by more than three times the upper limit of normal, or QTc prolongation on 12-lead electrocardiogram.

Bacterial DNA extraction

The reducing agent dithiothreitol (DTT [AppliChem, Darmstad, Germany], final concentration up to 5% for 15 min at room temperature) was used to homogenise the mucus phase of thawed sputum samples. Bacterial DNA from sputum and oropharyngeal (OPS) samples was then extracted using the DNeasy UltraClean microbial kit (Qiagen, Hilden, Germany), modified by pre-incubating with 9000 U Ready-Lyse lysozyme (Epicentre, Hessisch Oldendorf, Germany) for 1 hour at 37°C. Purified DNA was eluted in 30 μl of microbial DNA-free water (Qiagen).

16S rRNA amplicon quantification

The copy numbers of the 16S rRNA gene were determined by qPCR using previously reported primers specific to pan bacteria^1^ (see Supplementary Table E00). Amplification was performed using SsoAdvanced Universal SYBR Green Supermix (Bio-Rad, Hercules, CA) on a CFX96 Real-Time detection system (Bio-Rad) with the following cycling parameters: initial 2 min denaturation at 98 °C, followed by 45 cycles of 5 s denaturation at 98 °C, and 60 s annealing/elongation at 61.5 °C. Absolute quantification was performed based on a standard curve obtained with a purified amplicon product.

16S rRNA amplicon sequencing

Bacterial community composition was assessed by Illumina MiSeq sequencing with barcoded primers targeting the V1-V2 region (**Supplementary Table E00**). Amplification was performed using the Accuprime Taq DNA Polymerase High Fidelity kit (Invitrogen, Waltham, MA). Duplicate PCR reactions of 20 μl consisted of 2 μl of 10× Accuprime buffer II, 0.44 μl of each 10 mM barcoded primer F-27 and R-338, 9.03 μl of ultrapure water, 0.09 μl of AccuPrime Taq DNA Polymerase and 8 μl of DNA template with the following cycling parameters: initial 3 min denaturation at 94 °C, followed by 40 cycles of 30 s denaturation at 94 °C, 30 s annealing at 56 °C and 90 s elongation at 72 °C, with a final extension at 72 °C for 5 min. No-template PCR reaction controls (n = 2) were included. Amplicons were quantified using a LabChip GX instrument with the DNA 1 K kit (Perkin Elmer, Waltham, MA), pooled into equimolar amounts and purified using the AMPure XP bead cleaning system (Beckman Coulter, Brea, CA). Libraries were then diluted to 12 pM and spiked with 25% phiX before being loaded onto the Illumina MiSeq platform using pairwise chemistry, generating 250 × 2 read lengths.

Analysis of antibiotic resistance gene carriage

Quantification of carriage of antibiotic resistance genes (ARG) targeting 23S ribosomal RNA methyltransferases (*erm*(B) and *erm*(F)), ATP-binding cassette ribosomal protection protein (*mel* and *msr*[E]), major facilitator superfamily antibiotic efflux pump (*mef*), and tetracycline-resistant ribosomal protection proteins (*tet*[M] and *tet*[W]) was performed on sputum specimens using dye-based (SsoAdvanced Universal SYBR Green, Bio-Rad) or probe-based real-time PCR assays, using primer pairs, probes and conditions previously described^2^. To accommodate the limited material available for OPS samples, only *mel* and *tet*(W) gene expression was quantified, using the same qPCR protocol. Absolute quantification was performed based on standard curves obtained with purified amplicon products. The absolute copy number of resistance genes per sample was normalised to the 16S rRNA gene copy number used as a proxy for bacterial number. To obtain a synthetic picture of ARG carriage per sample, the absolute counts obtained for each individual gene were scaled from 0 to 1 to give equal importance to each gene, and the cumulative counts were reported.

Bioinformatics and statistical analysis

All analyses were performed in R version 4.1.0. Bioinformatics processing, which included demultiplexing, removal of chimeric and short reads, single-base resolution of reads into amplicon sequence variants (ASVs) using the Divisive Amplicon Denoising Algorithm 2 (DADA2) algorithm^3^ and taxonomic annotation using the SILVA database^4^, was performed using a dedicated pipeline available at <https://github.com/chuvpne/dada2-pipeline>.

Initial abundance filtering of absolute read counts (threshold > 1) was applied in phyloseq 1.38.0, reducing the total number of ASVs detected in all OPS and sputum samples, as well as in controls, from 4,233 to 4,170. Filtering based on ASVs belonging to the Bacteria kingdom further reduced the total number of ASVs from 4,170 to 4,099. Rank abundance analysis of the 15 most abundant ASVs in either patient samples or controls then identified 2 ASVs that were dominant only in controls (ASV6_*Cutibacterium*; ASV26_*Pseudomonas*) and were therefore excluded (**Supplemental Figure E5**). Additional sample filtering was applied based on absolute number of reads not exceeding that of the controls, which led to the exclusion of 10 out of 89 OPS samples and reduced the number of ASVs from 4,097 to 3,956 (**Supplemental Figure E4**).

For downstream analyses, a rarefied dataset with a read depth of 11,741 was used (**Supplemental Figure E2**), in which the relative abundance of each ASV was Hellinger transformed^5^ using the "decostand" function in vegan 2.6-2. The different alpha diversity measurements were performed using the alpha function in Microbiome 1.16.0. Principal coordinate analysis (PCoA) was used to visualise beta diversity based on unweighted UniFrac distance in vegan 2.6-2 and ggordiplots 0.4.1. Canonical correlation analysis (CCA) was performed using the cca function in vegan 2.6-2 to show the link between variation in respiratory microbiota composition and AR gene carriage. To compare changes in respiratory microbiota between the start and end of AZT treatment in patients with stable vs. increased ARG carriage, we performed a double PCoA, which combines analysis of phylogenetic and abundance data. To this end, we agglomerated the dataset at genus level, retained the 15 most abundant genera, and constructed a phylogenetic tree using the rtree function in ape 5.6-2. DPCoA was then performed using the ordinate function in phyloseq 1.38.0.

**Supplemental Figures**

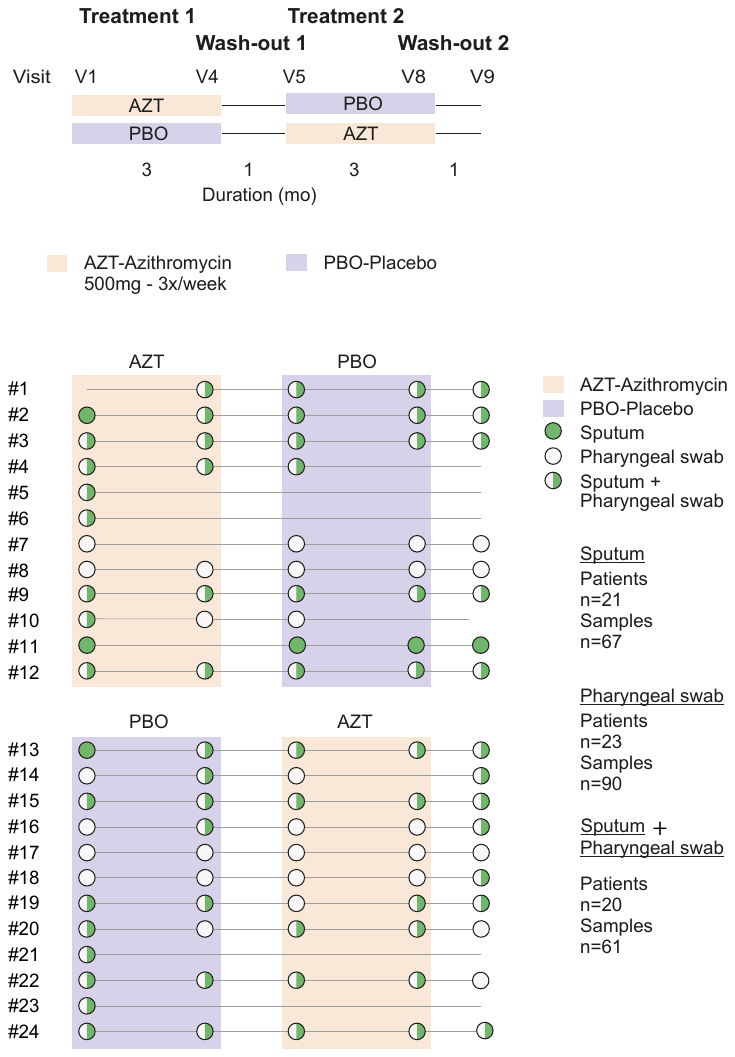

**Supplemental Figure E1**. Study design and specimen collection

| **A** | **B** |
| --- | --- |
| 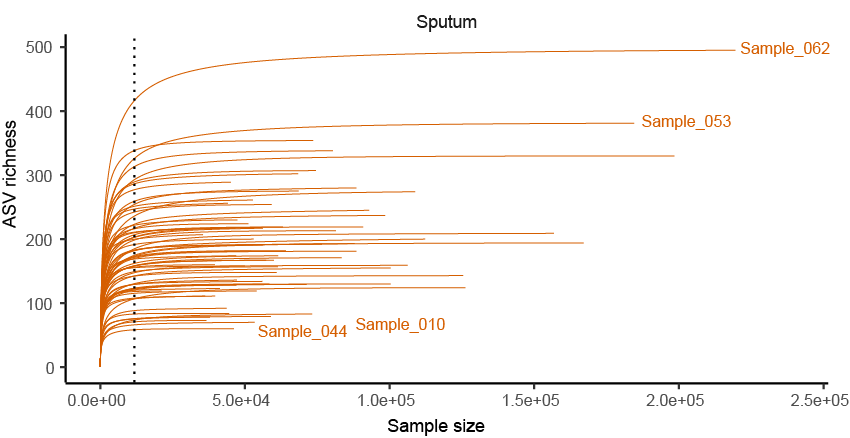 | 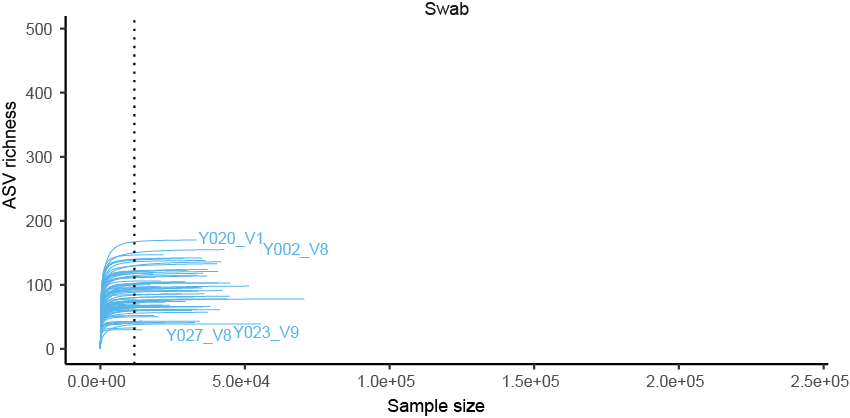 |

**Supplemental Figure E2.** Rarefaction. After abundance ﬁltering, a rarefied dataset in sputum (*A*) and OPS (*B*) samples, with a read depth of 11'741 was used for downstream analyses.

*Abbreviations*: OPS = oropharyngeal swab

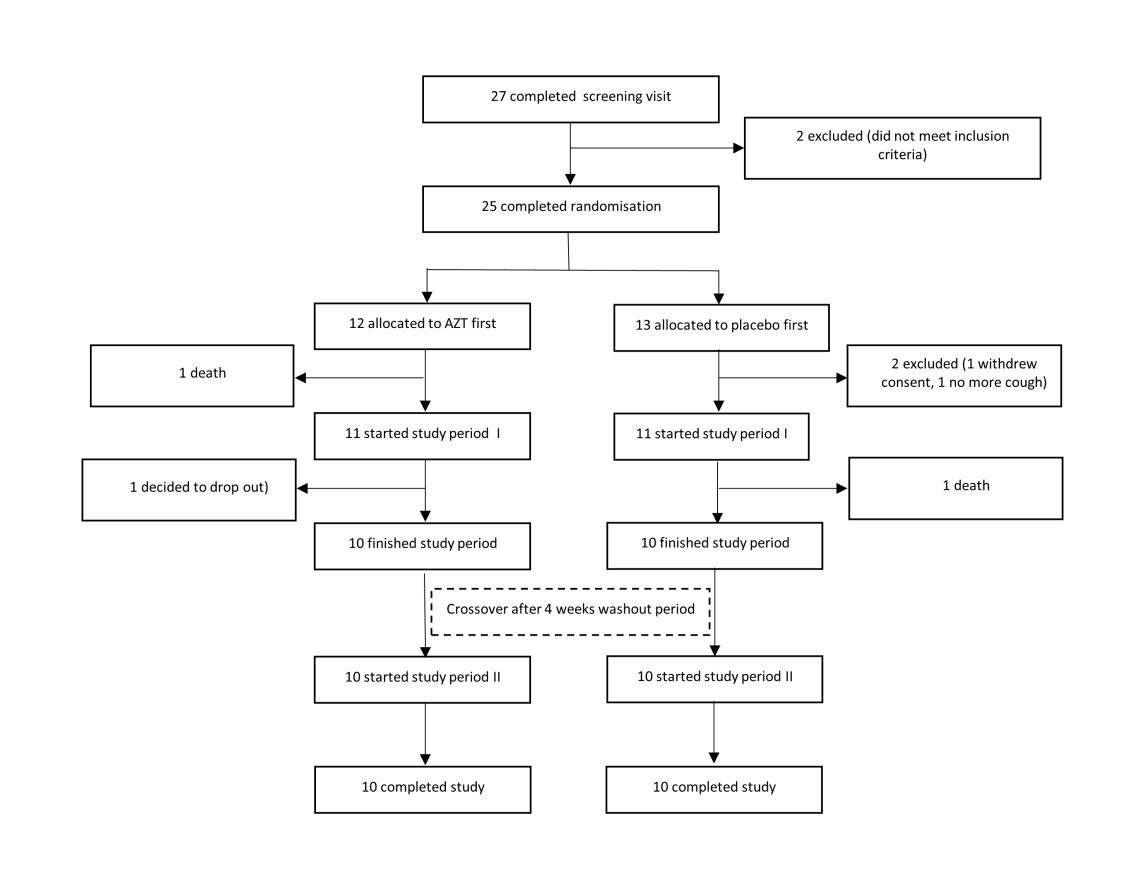

**Supplemental Figure E3.** CONSORT flow diagram of patients screening, inclusion, and analysis. The “*Azithromycin for the Treatment of Chronic Cough in Idiopathic Pulmonary Fibrosis*” was a prospective, randomized controlled crossover trial, of which the current study is a sub-analysis.

Threshold arbitrarily set at 1.1 x 10^4^ reads

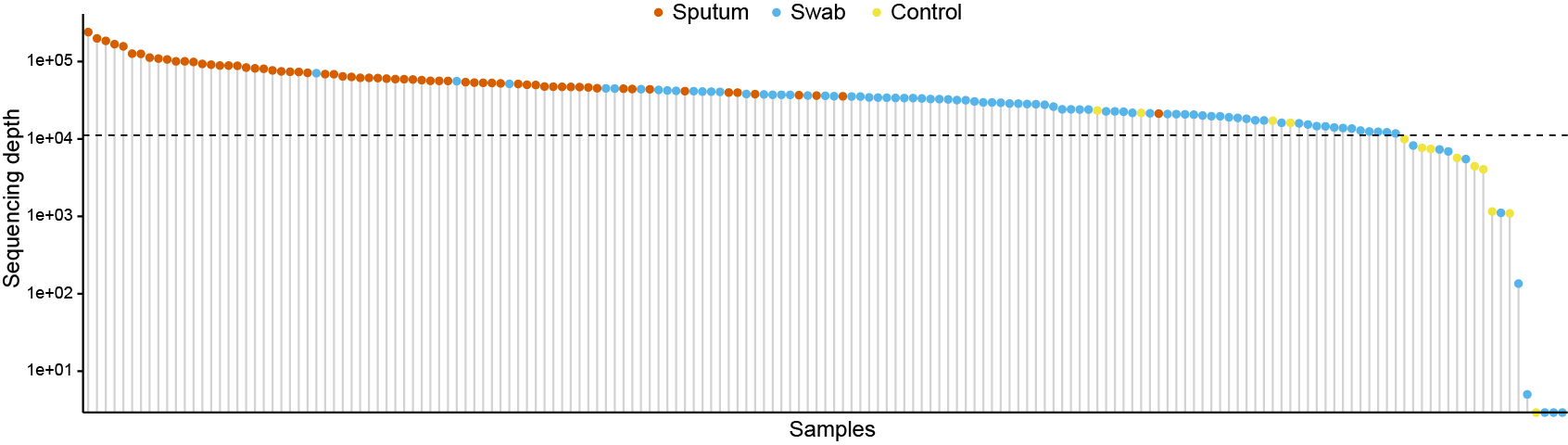
**Supplemental Figure E4.** Number of reads for each sputum, OPS and control sample. The threshold was arbitrarily set at 1.1 x 10^4^ reads. 10 OPS samples were excluded for further analysis.

*Abbreviations*: OPS = oropharyngeal swab

| **a)** | **b)** |
| --- | --- |
| 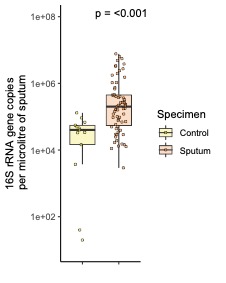 | 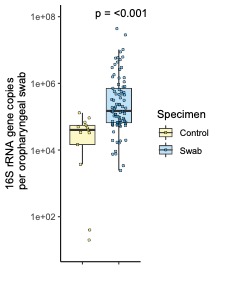 |

| **c)** | |
| --- | --- |
| 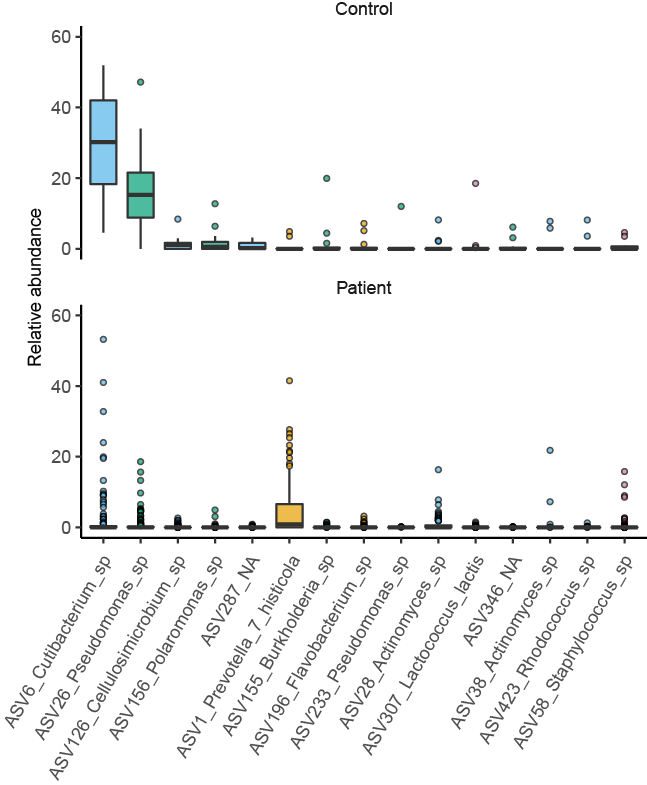 | 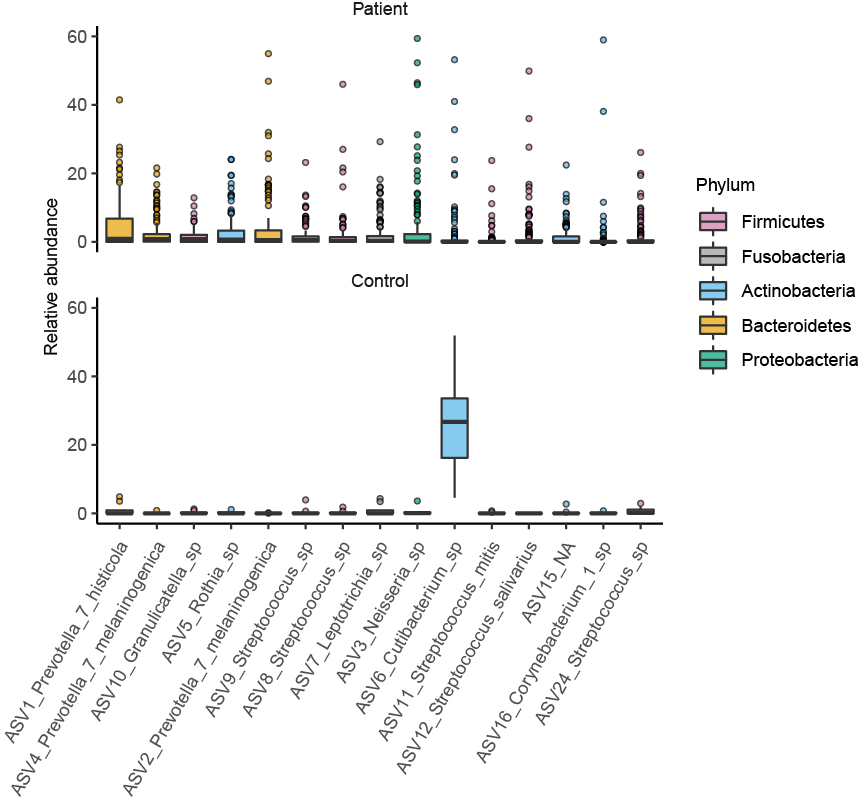 |

**Supplemental Figure E5.** Evidence of a distinct bacterial signal between patient samples and negative procedural controls. (*a* and *b*) The bacterial burden of sputum (*a*) and OPS (*b*) samples from patients was significantly greater than that of negative controls. The Wilcoxon rank sum test was used to compare the medians. (*c*) Rank abundance comparison of prominent taxa detected in negative control (left) and patient (sputum combined with OPS, right) samples. For each comparison, the 15 most abundant taxa in each group are displayed in decreasing order of median relative abundance.

*Abbreviations*: OPS = oropharyngeal swab

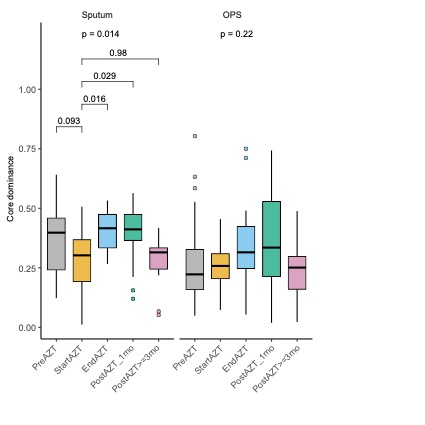

**Supplemental Figure E6.** Changes in community dominance during AZT treatment. Analysis of the dominance of the core community (species exceeding 0.2% relative abundance in more than 50% of the samples) showing a transient increase between the start and end of treatment in LRT, with only a similar trend in URT:

*Abbreviations*: AZT = azithromycin; LRT = lower respiratory tract; URT = upper respiratory tract

| **a)** | **b)** |
| --- | --- |
| 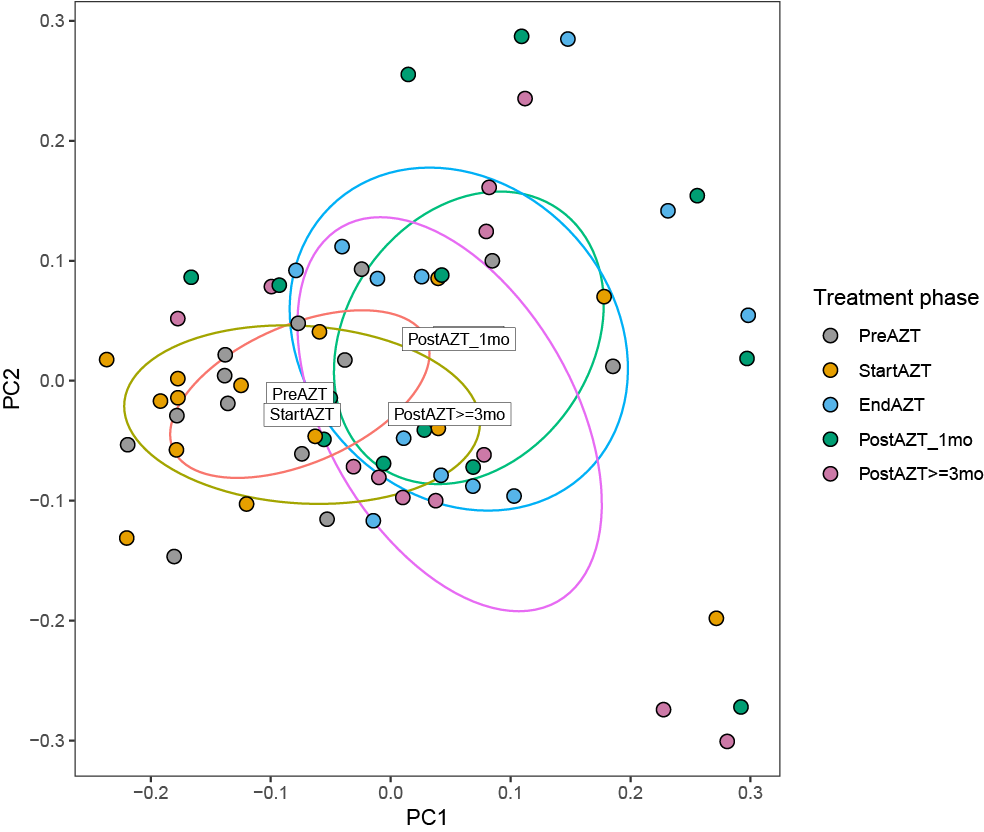  p = 0.008 | 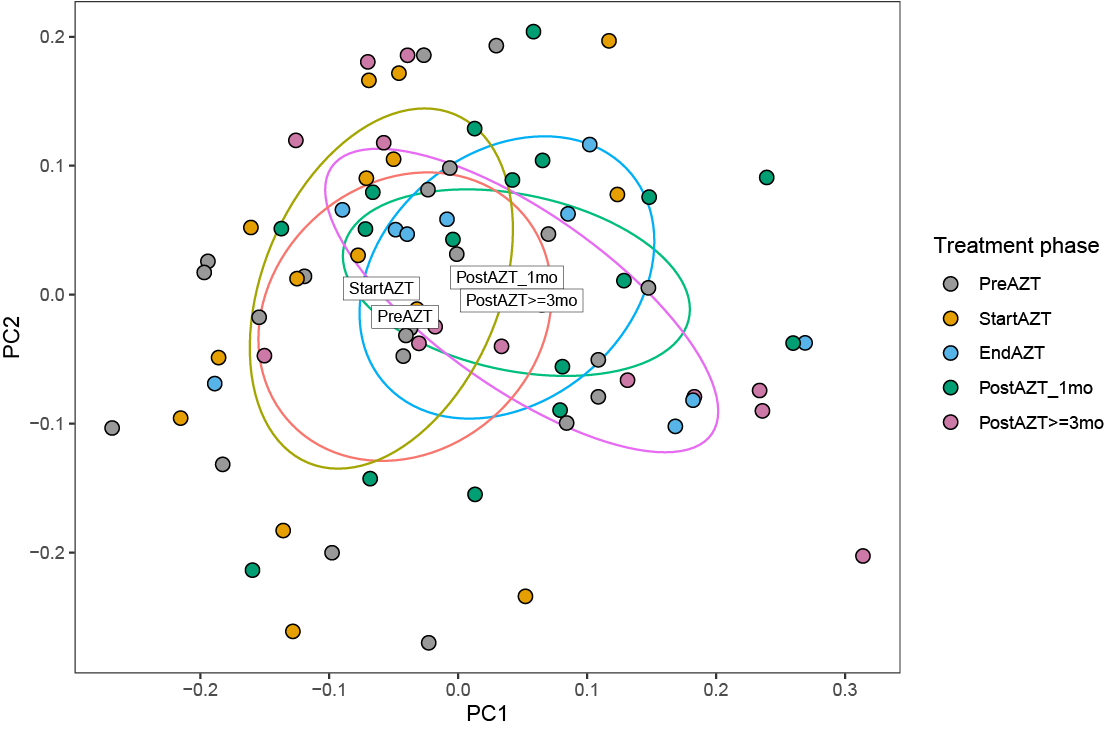  p = 0.009 |

**Supplemental Figure E7** (*A* and *B*) PCoA of bacterial communities based on unweighted UniFrac distance showing that the community composition of airway bacteria was distinct between specimens collected before or at the start of AZT treatment, compared to those collected at the end of treatment or later, in LRT (*a*) and URT (*b*). In PCoA labels indicate the position of the centroid of the corresponding group, the label “EndAZT” is hidden by the label “PostAZT_1mo”.

*Abbreviations*: PCoA = principal coordinate analysis; AZT = azithromycin; LRT = lower respiratory tract; URT = upper respiratory tract

| **a)** | **b)** |
| --- | --- |
| 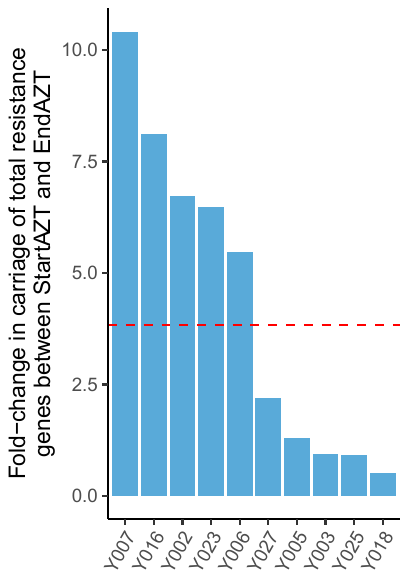 | 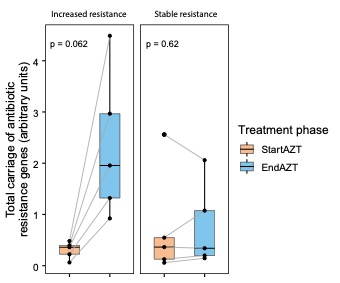 |

**Supplemental Figure E8.** Patient-specific fold-change in the cumulative sum of seven pooled ARG in the LRT during AZT treatment. (*a*) Separation into two groups of patients according to the median fold-change (3.8; red dotted line; ranges 5.5-10.4 and 0.51-2.2 for the groups of patients considered to have shown increased resistance, respectively stable resistance, during treatment). (*b*) Kinetics of ARG carriage in the two groups of patients with increased or stable resistance during AZT treatment.

*Abbreviations*: ARG = antibiotic resistance genes; LRT = lower respiratory tract; AZT = azithromycin

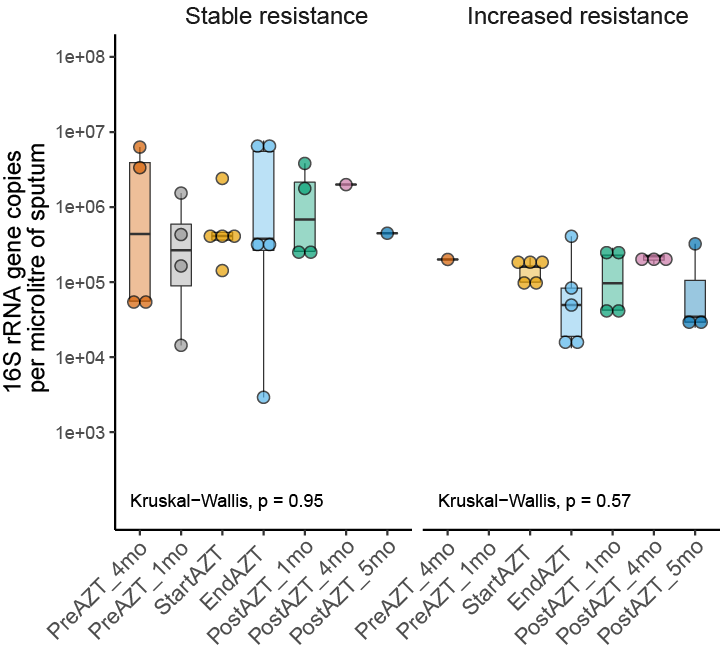

**Supplemental Figure E9.** Bacterial load evolution between treatment phases in LRT. (*A*) There was no change in bacterial load between treatment phases, regardless of ARG carriage.

*Abbreviations*: LRT = lower respiratory tract; ARG = antibiotic resistance genes

| **a)** | **b)** |
| --- | --- |
| 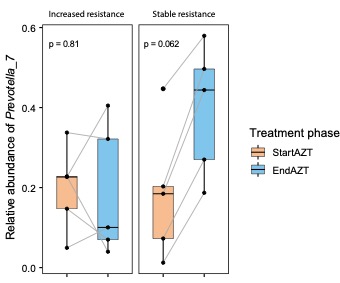 | 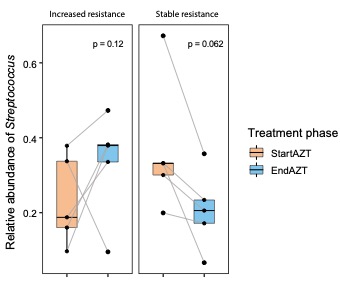 |
| **c)** | **d)** |
| 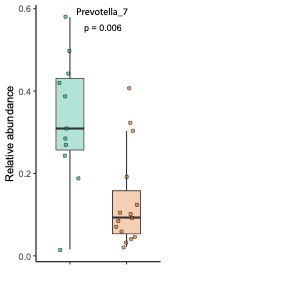 | 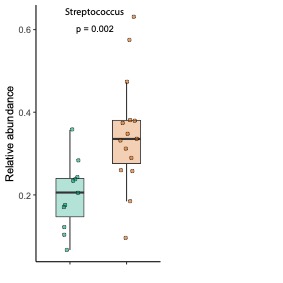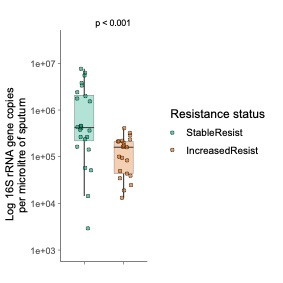 |

**Supplemental Figure E10.** Changes in the composition of the LRT microbiota in relation to ARG carriage during AZT treatment. (*a*) Increase in the relative abundance of the genus *Prevotella_7* in all five patients with stable ARG carriage and only one patient with increased ARG carriage. (*b*) Increase in the relative abundance of the genus *Streptococcus* in four patients with increased ARG carriage. (*c*) Relative abundance of *Prevotella_7* showing higher levels in patients with stable resistance. (*d*) Relative abundance of *Streptococcus* showing higher levels in patients with increased resistance.

*Abbreviations*: LRT = lower respiratory tract; ARG = antibiotic resistance genes; AZT = azithromycin

| **a)** | **b)** |
| --- | --- |
| 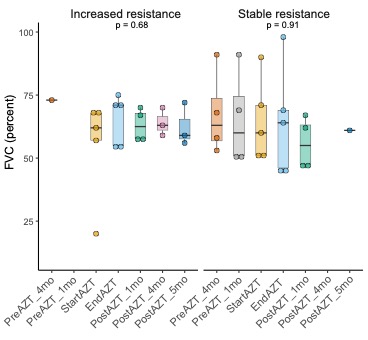 | 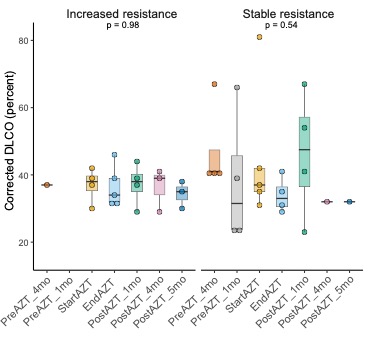 |

**Supplemental Figure E11.** Links between AZT treatment-related carriage of ARG in the LRT microbiota, lung function and oxygen therapy. (*a* and *b*) Analysis of LRT samples from patients with available ARG carriage status (n = 10) showing the absence of variations in FVC (*a*) and DLCOc (*b*) during the different phases of treatment, independently of AR gene carriage.

*Abbreviations*: AZT = azithromycin; ARG = antibiotic resistance genes; LRT = lower respiratory tract; FVC = forced vital capacity; DLCOc = corrected diffusing capacity of the lung for carbon monoxide

**Supplemental Table**

| **Table E1.** Oligonucleotide primers for 16S ribosomal RNA gene analysis. | | |
| --- | --- | --- |
| **Name** | **Method** | **Sequence (5’-3’)** |
| 27F | Illumina MiSeq | **AATGATACGGCGACCACCGAGATCTACAC***TATGGTAATTCC*AGMGTTYGATYMTGGCTCAG |
| 338R | Illumina MiSeq | **CAAGCAGAAGACGGCATACGAGAT**NNNNNNNNNNNN*AGTCAGTCAGAA*GCTGCCTCCCGTAGGAGT |
| 926F | qPCR | AAACTCAAAKGAATTGACGG |
| 1062R | qPCR | CTCACRRCACGAGCTGAC |
| Note: 27F and 338R Illumina sequencing primers allow to amplify the V1-V2 hypervariable region of the 16S rRNA gene. Primers 926F and 1062R target conserved sequences flanking the V6 hypervariable region of the 16S rRNA gene (Bacchetti de Gregoris, doi: 10.1016/j.mimet.2011.06.010). Boldface indicates Illumina adapter sequences, italicized characters indicate linkers, and underlined characters indicate sequences annealing to the target gene. The sequence NNNNNNNNNNN represents the sample-specific molecular identification barcode. | | |
